# Estimation of a state of Corona 19 epidemic in August 2020 by multistage logistic model: a case of EU, USA, and World (Update September 2020)

**DOI:** 10.1101/2020.08.31.20185165

**Authors:** Milan Batista

## Abstract

The article provides an estimate of the size and duration of the Covid-19 epidemic in August 2020 and September 2020 for the European Union (EU), the United States (US), and the World using a multistage logistical epidemiological model.

## Introduction

This article discusses the Corona 19 epidemic that outbreaks in Wuhan (China) in December 2019. At the outbreak, the epidemic final size and its duration are common questions. (Brauer, 2019a, 2019b; Fisman D, 2014; Hethcote, 2000; House, Ross, & Sirl, 2013). Different models are used to answer such a question. These include models based on logistic function or Richards function (Batista, 2020a; Pongkitivanichkul et al., 2020; Roberts, 2020; Zou Y et al., 2020), deterministic classical and enhanced SIR and SEIR models (Anastassopoulou, Russo, Tsakris, & Siettos, 2020; Giordano et al., 2020; S. B. He, Peng, & Sun, 2020; Loli Piccolomini & Zama, 2020; Lopez & Rodo, 2020; Maier & Brockmann, 2020; Ming, Huang, & Zhang, 2020; Nesteruk, 2020; Peng, Yang, Zhang, Zhuge, & Hong, 2020; Tang et al., 2020; Wu, Leung, & Leung, 2020; C. Y. Yang & Wang, 2020), statistical-based models (S. He, Tang, & Rong, 2020; Mbuvha R & T, 2020; Roda, Varughese, Han, & Li, 2020; Verity, Okell, & Dorigatti, 2020; W. Yang, Zhang, Peng, Zhuge, & Hong, 2020; Zahiri, RafieeNasab, & Roohi, 2020; Zhan, Tse, Lai, Hao, & Su, 2020), time-series models (Agosto & Giudici, 2020; Ceylan, 2020), a new models (Nesterov, 2020; Singhal, Singh, Lall, & Joshi, 2020).

There are at least two problems with the modeling of the epidemic. First, the question is whether a chosen model is an appropriate description of the epidemic, especially if the epidemic has several separate outbreaks or is dragging into a new wave. The second is that at the beginning of the outbreak or at a new wave, the parameters of the models are not known (Keeling & Rohani, 2008), or better they depend on the course of the epidemic. Therefore, prediction using such models are unlikely to be successful or should be used with caution, especially if used for long-term forecasting. However, when a model is a reasonable description of the epidemic, then the long term trend of an epidemic may be assessed by monitoring changes in the model parameters. It is clear that when the parameters of the model retain their values, a long-term prediction is possible because the epidemic curve is determined. We will call such an epidemic state stable; otherwise, the state is unstable. Here we stress that any new local outbreak or import of infected into the population can destabilize the situation; almost no model can predict this. The best that models can offer are solutions for selected scenarios that may or may not realize (Bettencourt & Ribeiro, 2008; Klepac, Kissler, & Gog, 2018). (Bettencourt & Ribeiro, 2008; Klepac, Kissler, & Gog, 2018).

In sequel will use a multistage logistic model to assess the state of the epidemic in EU, US and World. The model is not new. Two stages logistic model was used for modeling 2003 SARS outbreak in Toronto (Canada) (Hsieh & Cheng, 2006; Wang, Wu, & Yang, 2012), and the multistage logistic model was introduced by Chowell et (Chowell, Tariq, & Hyman, 2019) was used for modeling Spanish flu of 1918 in Genova, Switzerland (Chowell, Ammon, Hengartner, & Hyman, 2006). We note that a multistage model based on SEIR model was introduced by Abdulrahman (Abdulrahman, 2020).

The data used in this article are total confirmed cases up to 30 August 2020, as are daily reported by Worldmeter^1^. We do not enter into the question of how good and reliable these data are.

In the Appendix assessment of the epidemic on the basis of data by the end of September 2020.

### The model

The base of the multistage (or multi-wave) logistic model is the logistic model, which is also called a simple epidemic model (Bailey, 1975) or the SI (susceptible-infective) model (Frauenthal, 1980). The basic equation of the logistic model is (Daley & Gani, 2001; Frauenthal, 1980)

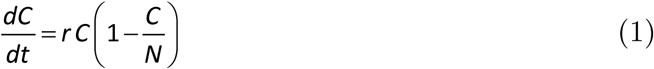

where *t* is the time, *C* (*t*) is the total number of cases in time *t, r* > 0 is infection rate, and *N* is the size of the susceptible population. If *C* (0) = *C*_0_ > 0 is the initial number of cases, then the solution of (1) is

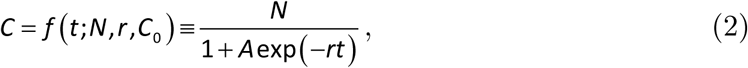

where 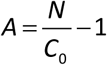.

Now, assuming that epidemic is composed of *n*_*w*_ mutually separated waves, then the model (2) can be generalized as follows

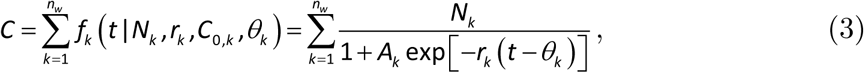

where 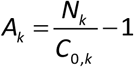. For each wave, we, therefore, have four unknown parameters: *N*_*k*_ > 0, *C*_*0,k*_ > 0, *r*_*k*_ > 0, and *θ*_*k*_ > 0 is the time delay, where for the first wave *θ*_1_ = 0. We did not incorporate parameter *A*_*k*_ into the exponential function because we want *θ*_*i*_ to be in the data time range. The total number of unknown parameters is therefore

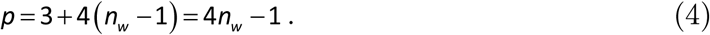

If the number of data are *n* then *n* > *p* and therefore the number of waves that can be detected is limited by

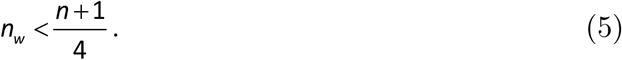

From (3), we can calculate the total size of the susceptible population

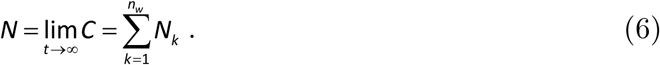

We follow Daley and Gani and define the end of the epidemic when the number of infectives is within 1 of its final size (Daley & Gani, 2001). Thus, to determine the end of the epidemic must be solved numerically (3) for *t* where we set *C* = *N* − 1,

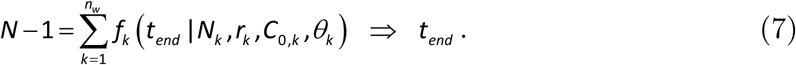

The above model was implemented in the Matlab program *fitVirusXX* (Batista, 2020b). In the program, the parameters of the model are estimated by the ordinary least-squares method by minimizing the following expression

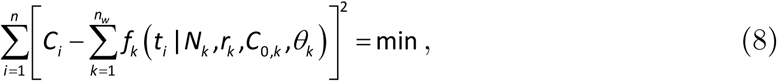

where *C*_1_, *C*_2_, …,*C*_*n*_ are the reported cases in times *t*_1_, *t*_2_, …,*t*_*n*_. The above function has many possible local minimum values. Therefore, a heuristic approach was used with the brute force search method to determine a quasi-minimum of (8).

## Results

### European Union

Figure 1 shows that the course of the epidemic in the EU so far can be described in three waves. EU countries introduced strict quarantine in March so that the first wave peaked in early April and then weakened by mid-June. Already in April, a second smaller wave appeared, but it did not have a pronounced peak; its effect was only that the first wave dragged on into June. After the release of the measures, a new summer wave began to rise in early July.

**Figure 1.**
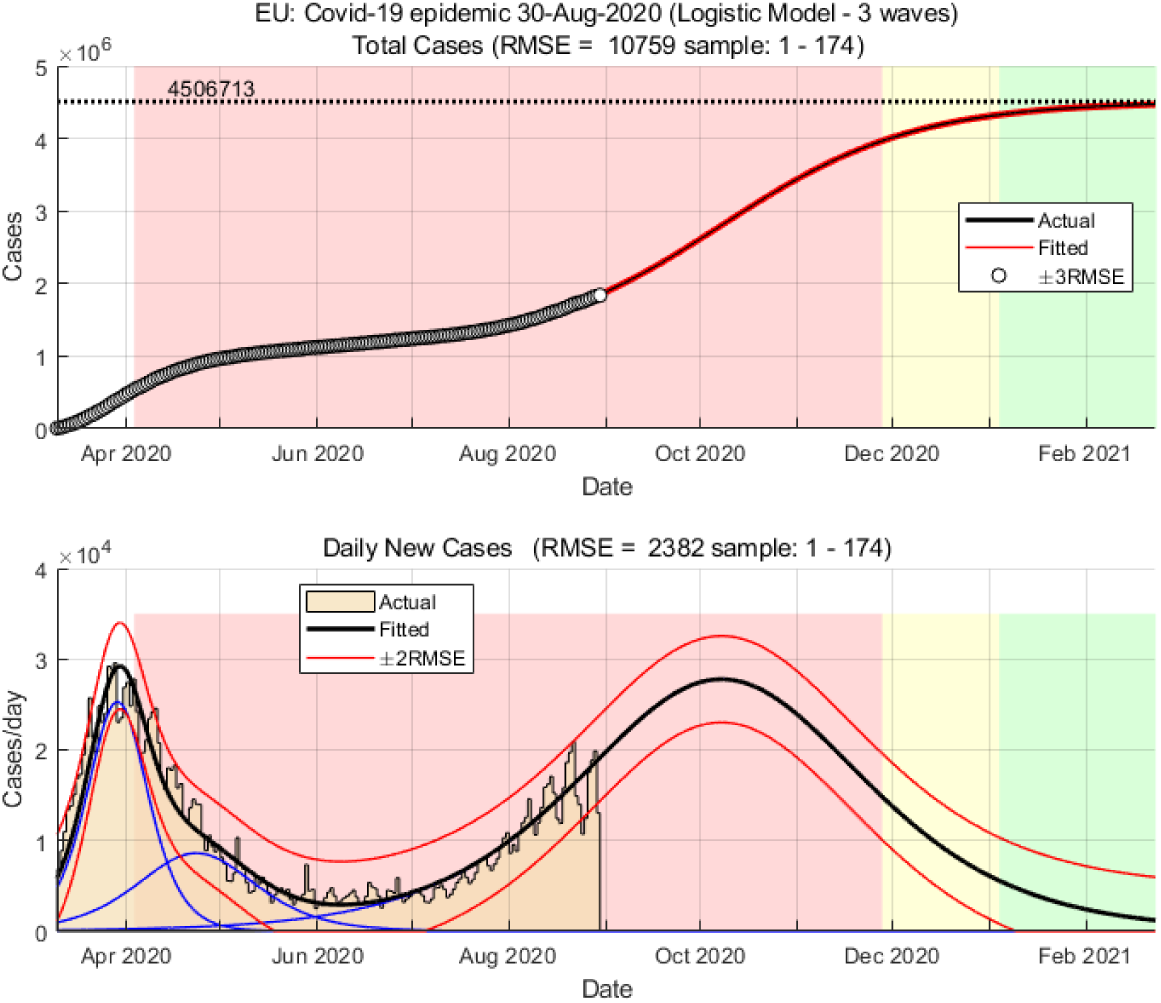
Covid-19 epidemic in EU – current state and prediction (red area 12-88% total cases, yellow area 89-96% total cases)

From the graph in Figure 2, we can see that the epidemic in the EU has so far shown no signs of calming down. The estimate of the final number of infections, as well as the duration of the epidemic, has been steadily increasing since April. In August, the course of the epidemic passed into a markedly unstable phase, where, as can be seen from Figure 2, no clear trend can be observed. The current estimate shows a final 3.8 to 4.5 million infections and a duration of 600 to 700 days, i.e., until the winter of 2022.

**Figure 2.**
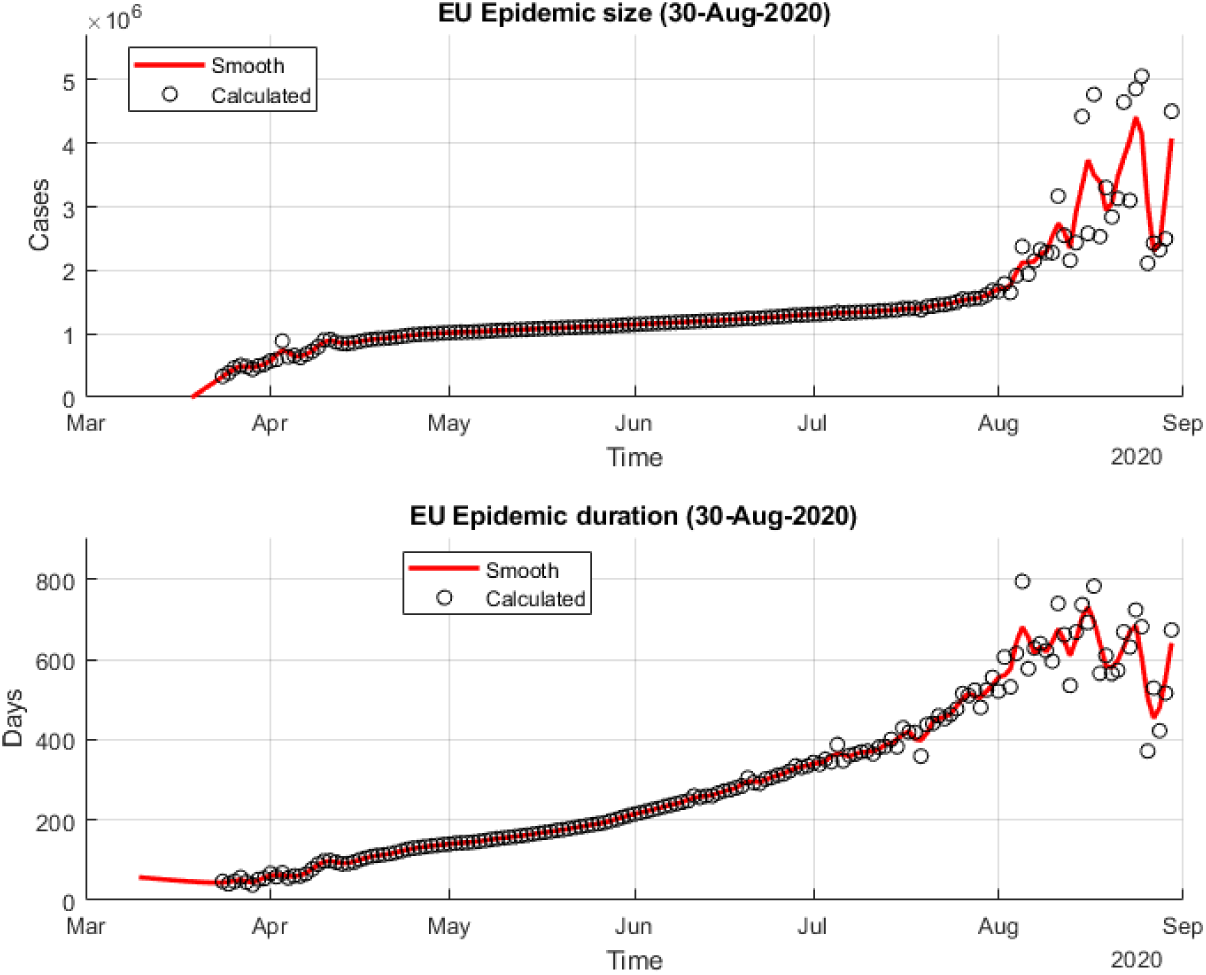
Daily-predicted epidemic size and duration for Covid 19 in EU.

**Figure 3.**
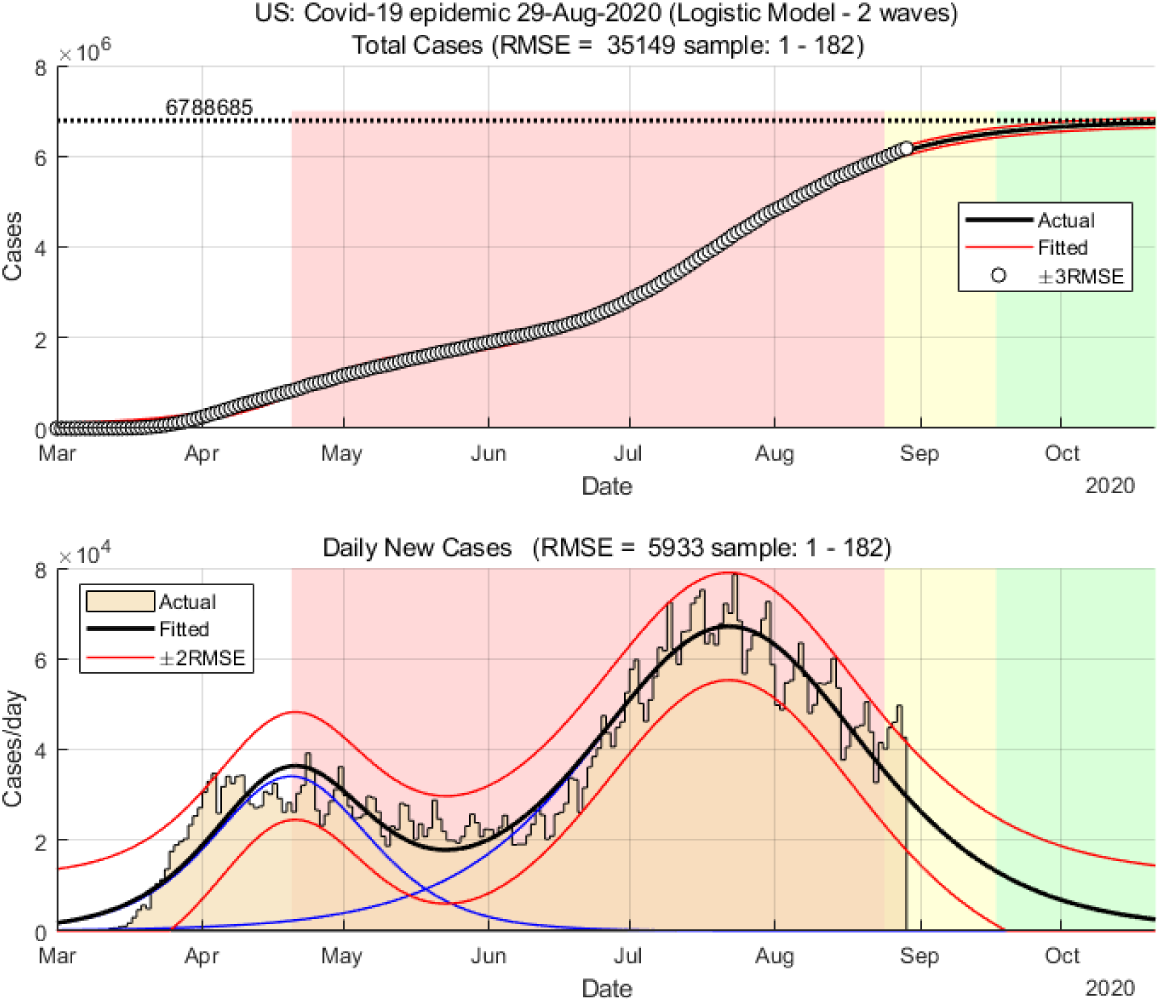
Current state and forecast of Covid 19 in USA.

### The United States

In Figure 2, we can see that the epidemic in the U.S. has two waves, the first smaller reaching its peak in early May and the second larger at the end of August. In the graph in Figure 4, we can see that the trend in predicting the size of the epidemic and its duration was linear, then began to rise sharply at the end of June and reached its peak in mid-June with an estimate of 10 million final infections. This was followed by an unstable period of declining size estimates, and in the last two weeks of August, this estimate stabilized at about 6.3 million total infections. The estimate of the duration of the epidemic stabilized at 434 days, i.e., the epidemic is expected to last until May 2021.

**Figure 4.**
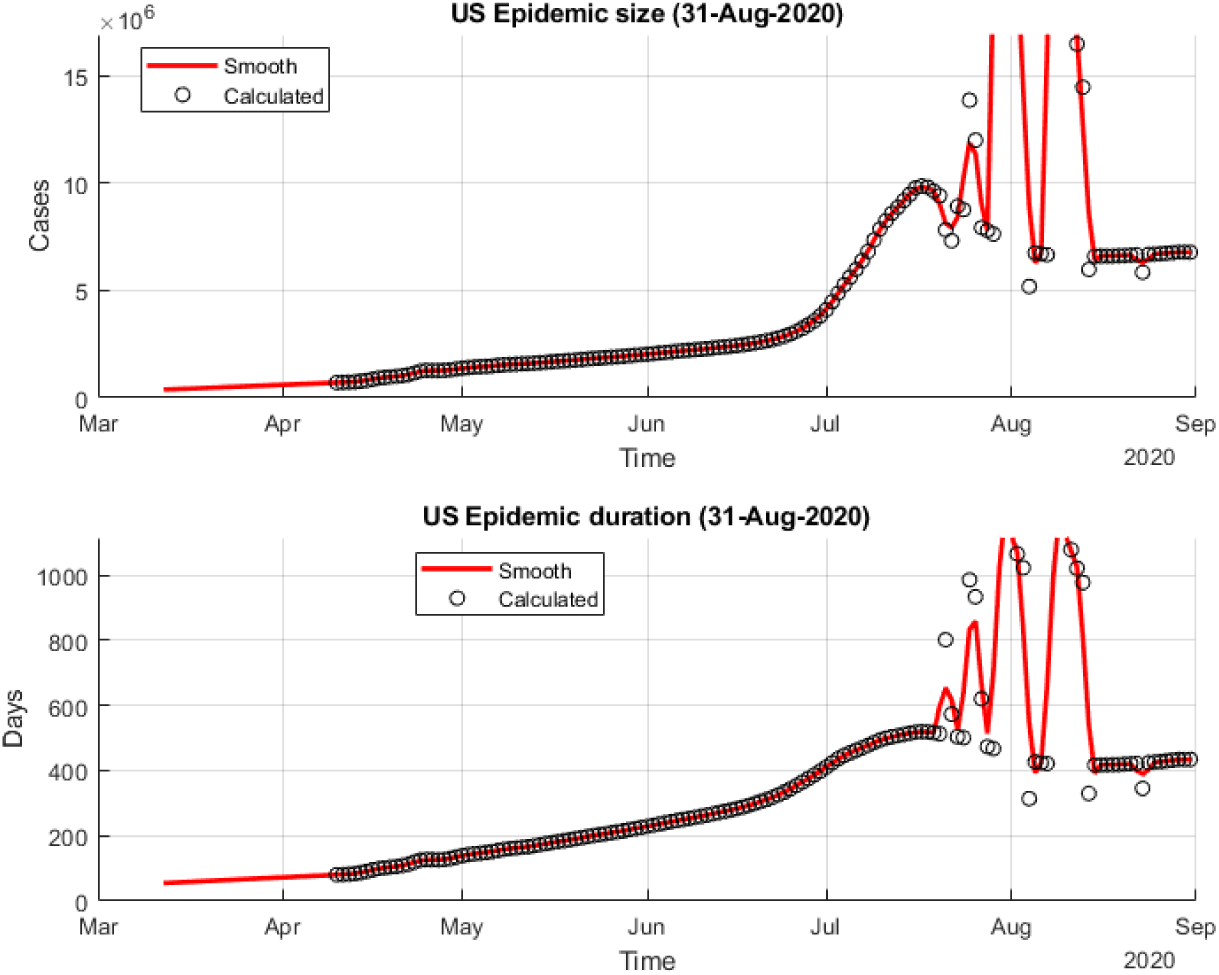
Daily-predicted epidemic size and duration for Covid 19 in US.

### World

Similar to the EU, we see in Figure 5 that the current course of the epidemic around the World can be divided into three waves. The first peaked in April, the second wave extended the epidemic in June, followed by a stronger third wave, which peaked in mid-August. From the graph in Figure 6, we can see that the course of the epidemic until the beginning of June was a steady increase in the estimation of its final size and duration. This was followed by a period of an indistinct but growing trend, which is not yet showing signs of calming down. The current estimate of the final size is 30 to 40 million infections and a duration of 450 d0 700 days, i.e., the epidemic could drag on into winter 2022.

**Figure 5.**
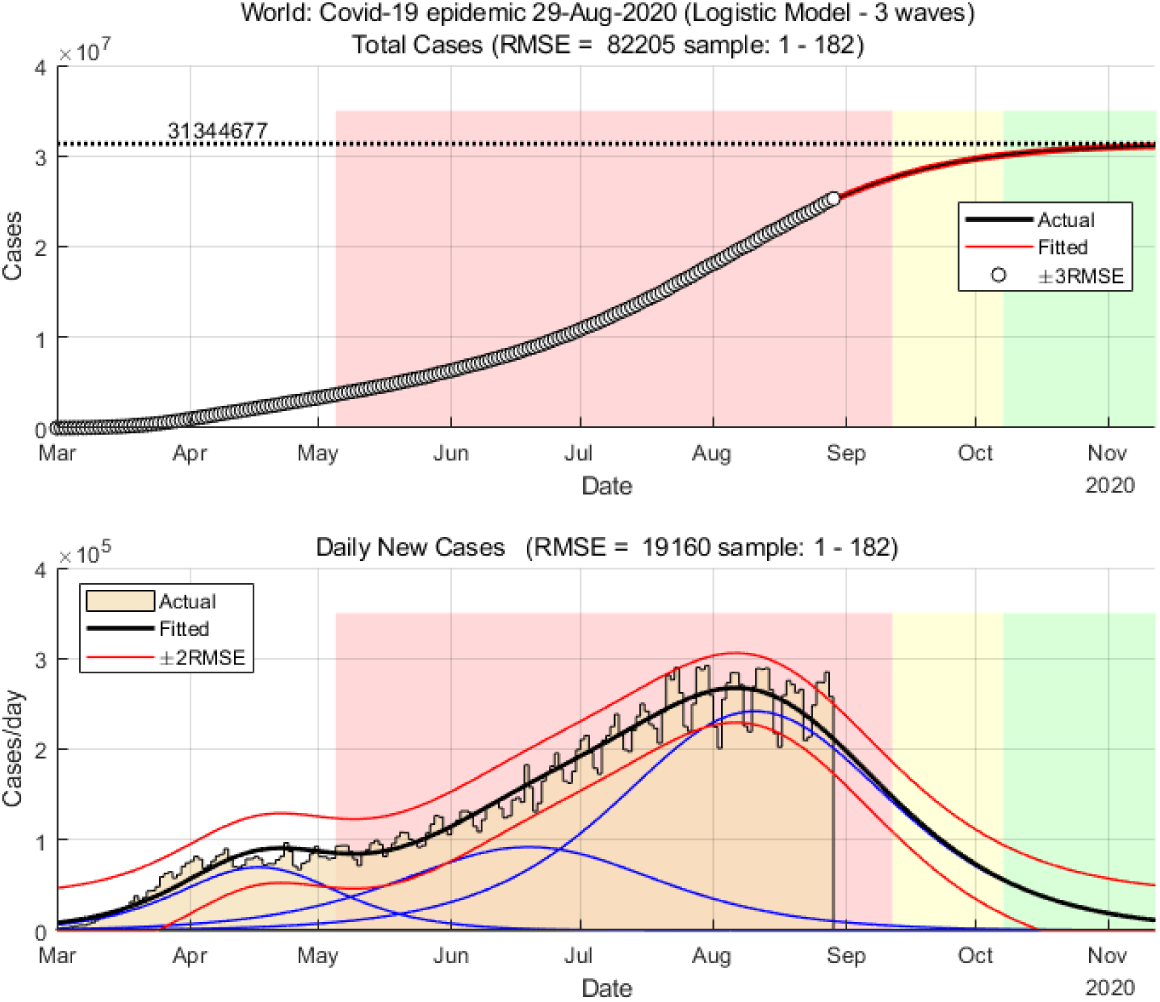
Situation and forecast of Corona 19 epidemic in the World.

**Figure 6.**
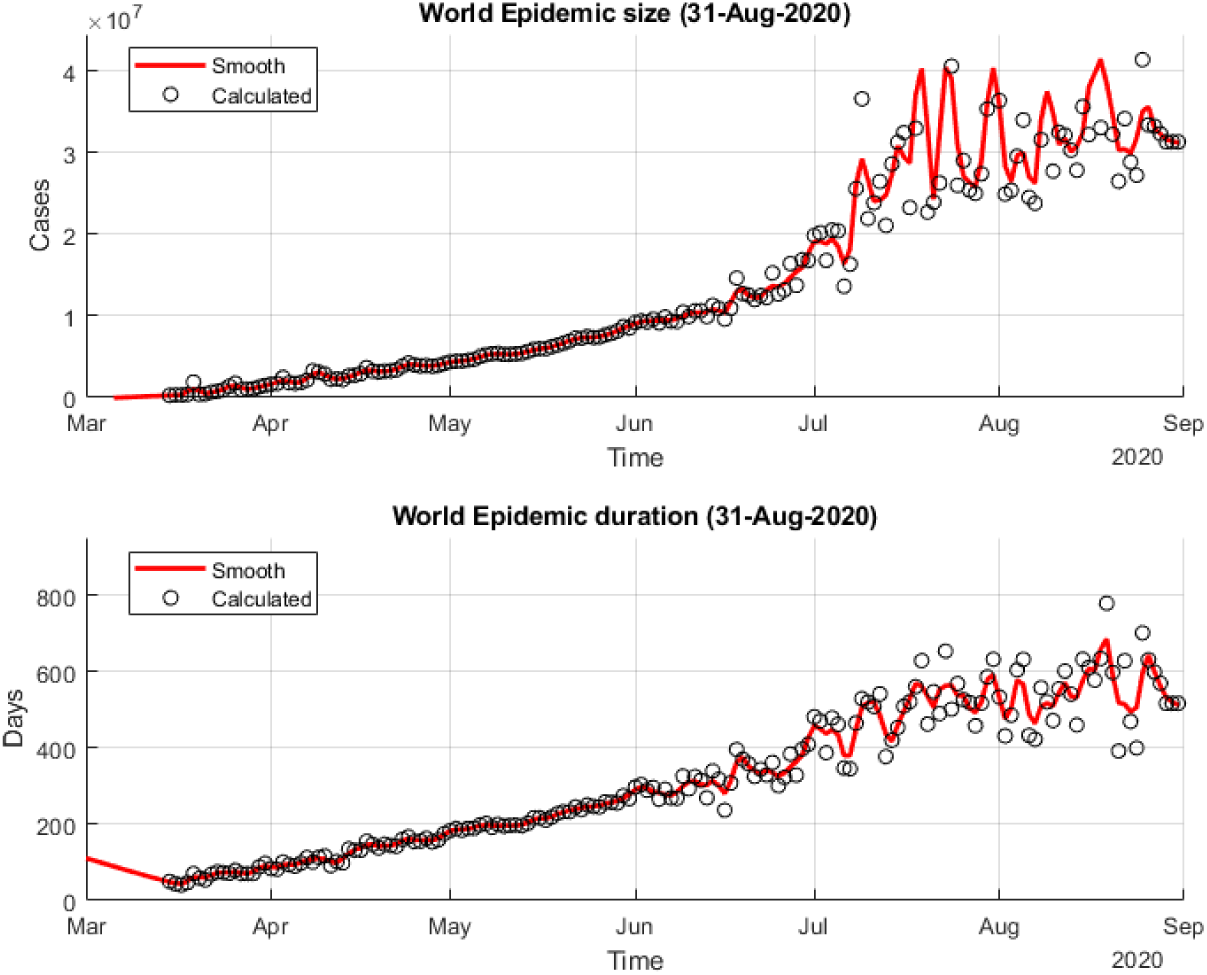
Daily-predicted epidemic size and duration for Covid 19 in the World.

## Conclusions

We first note that each case under consideration has a different course of estimates of the final size and duration of the epidemic. The absence of a unique pattern makes the prediction of the final size and duration of the epidemic difficult. Namely, even if a convergence of parameters is achieved, we cannot be sure that this is the last phase of the epidemic; a new outbreak is possible at any time.

For now, only parameters for the US achieve convergence. It can be estimated that about 6.5 million peoples will be infected, and the epidemic will last about 450 days, i.e., until May 2021, if, of course, no third wave will emerge.

A new wave of epidemics is rising in the EU, which is expected to peak in October. However, the daily estimates of the model parameters for the EU are extremely unstable, so that every forecast so far is questionable and will certainly change. Similarly, we can conclude about the course of the epidemic around the World. The epidemic has so far crossed the top of the third wave, but the situation is not yet stable.

In the end, we stress that these predictions are not final but only reflect the current data.

Appendix.

Assessment of the epidemic on the basis of data by the end of September 2020.

## European Union

The situation in the EU is still chaotic; i.e., the course of the epidemic does not yet have a clear trend. According to current data, the estimate of the size of the epidemic has risen to 18 million confirmed tests. The peak is expected to be reached sometime in late December when the daily number of confirmed tests would be just over 10,000. The end of the epidemic is expected to be sometime in late April 2021. But, as has been said, the situation is still chaotic, so these estimates are likely to change in the future.

**Figure 1a.**
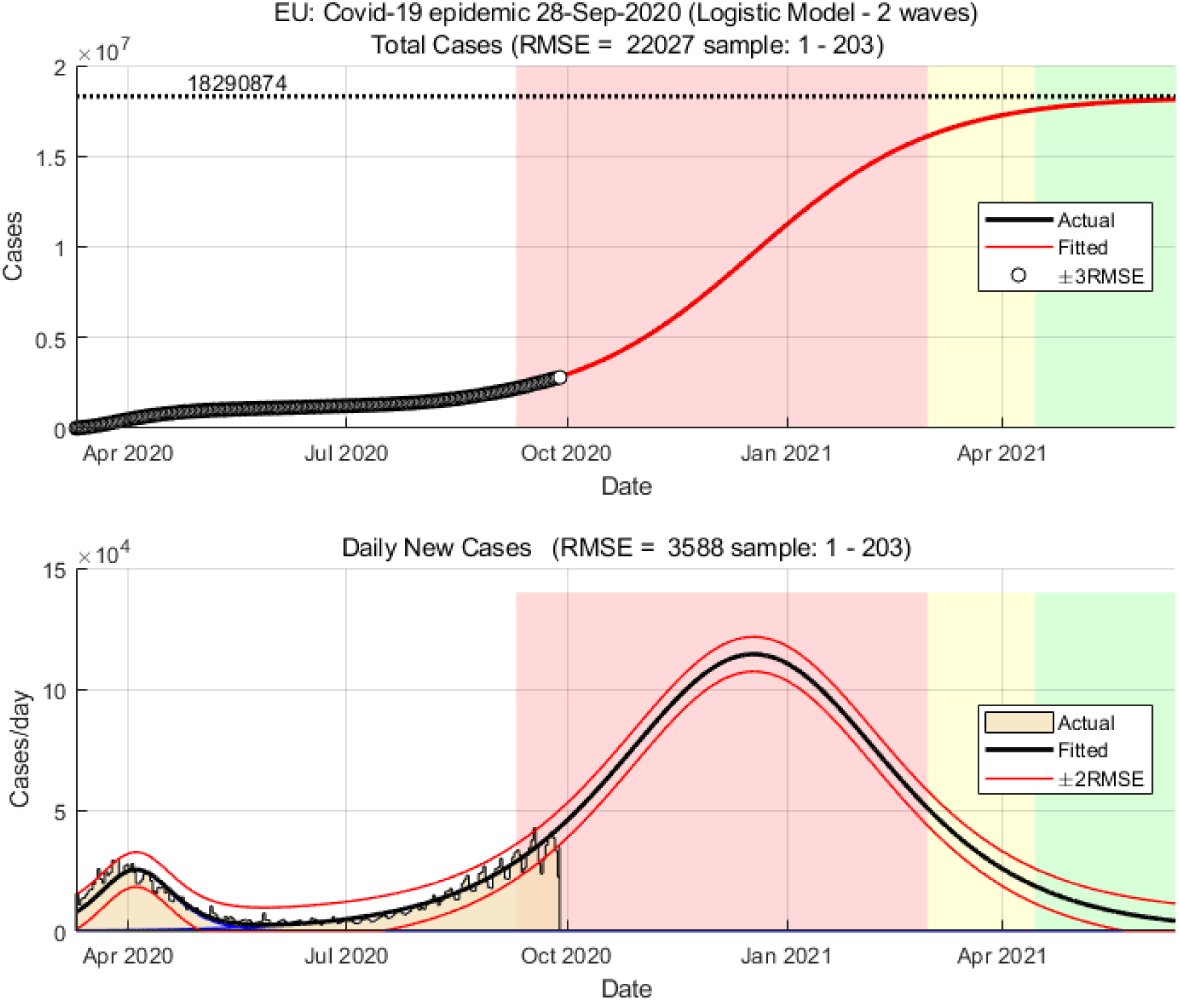
Covid-19 epidemic in EU – current state and prediction (red area 12-88% total cases, yellow area 89-96% total cases)

**Figure 2a.**
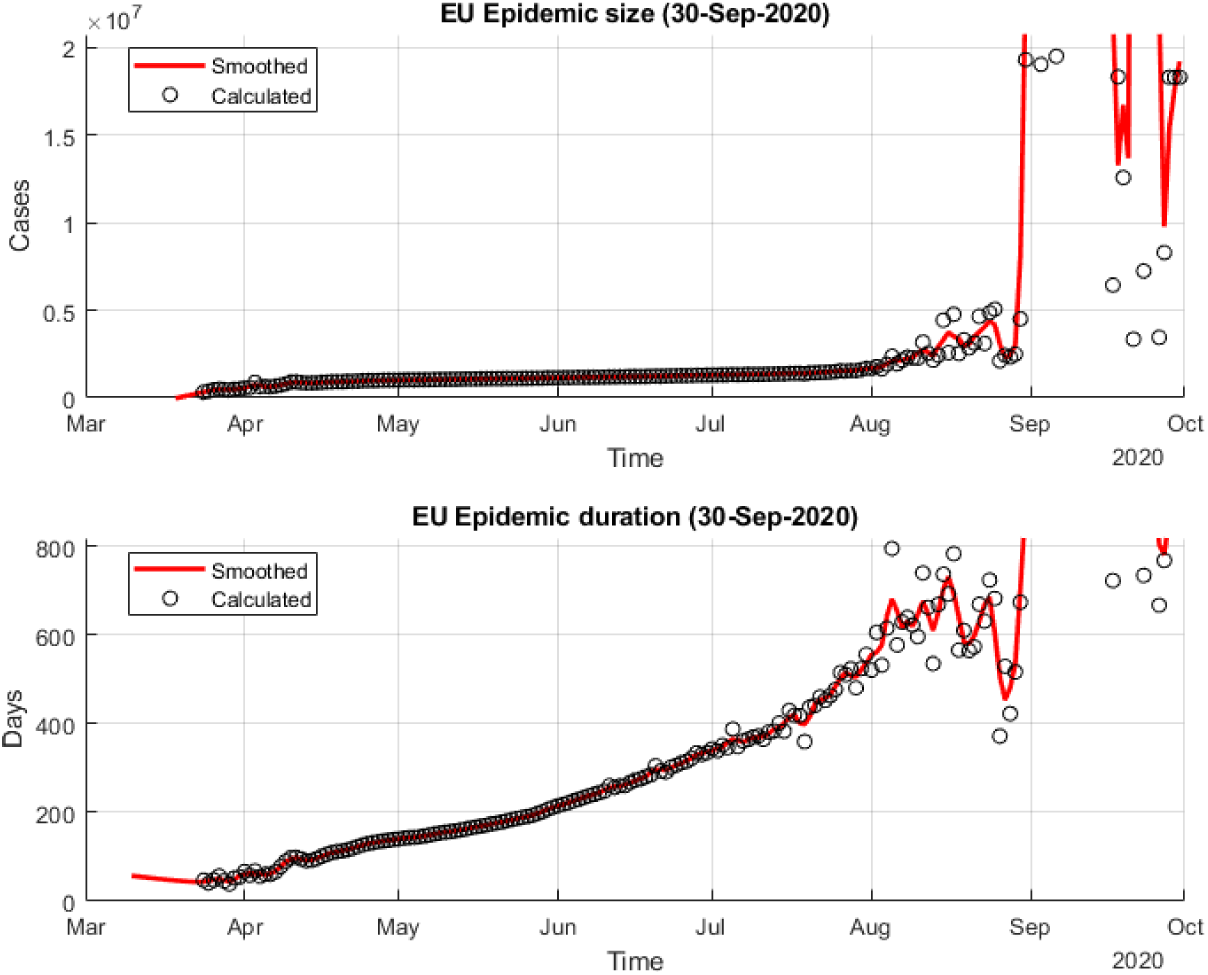
Daily-predicted epidemic size and duration for Covid 19 in the EU.

**Figure 3a.**
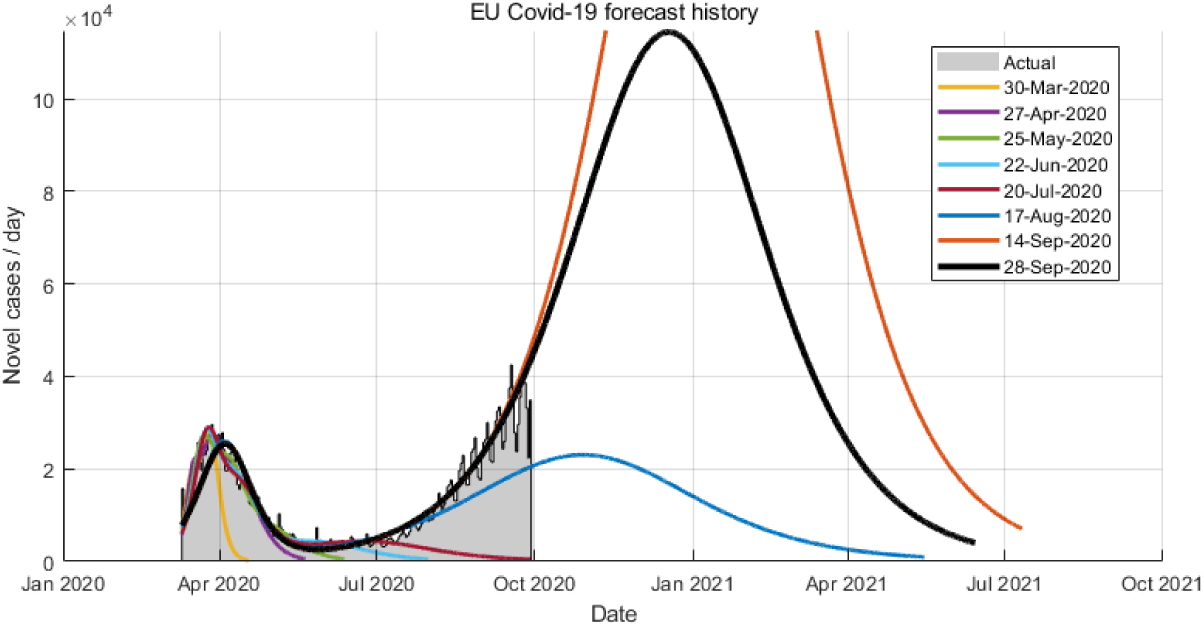
History of incidence forecasting for Covid 19 in the EU.

## The United States

The data suggest that a third wave could start in the US. The trend of the size of the epidemic and its duration are clear, but they do not yet show a clear end. The current estimate is that the wave could cover up to 9 million people; the third wave is expected to end sometime in early January 2021.

**Figure 4a.**
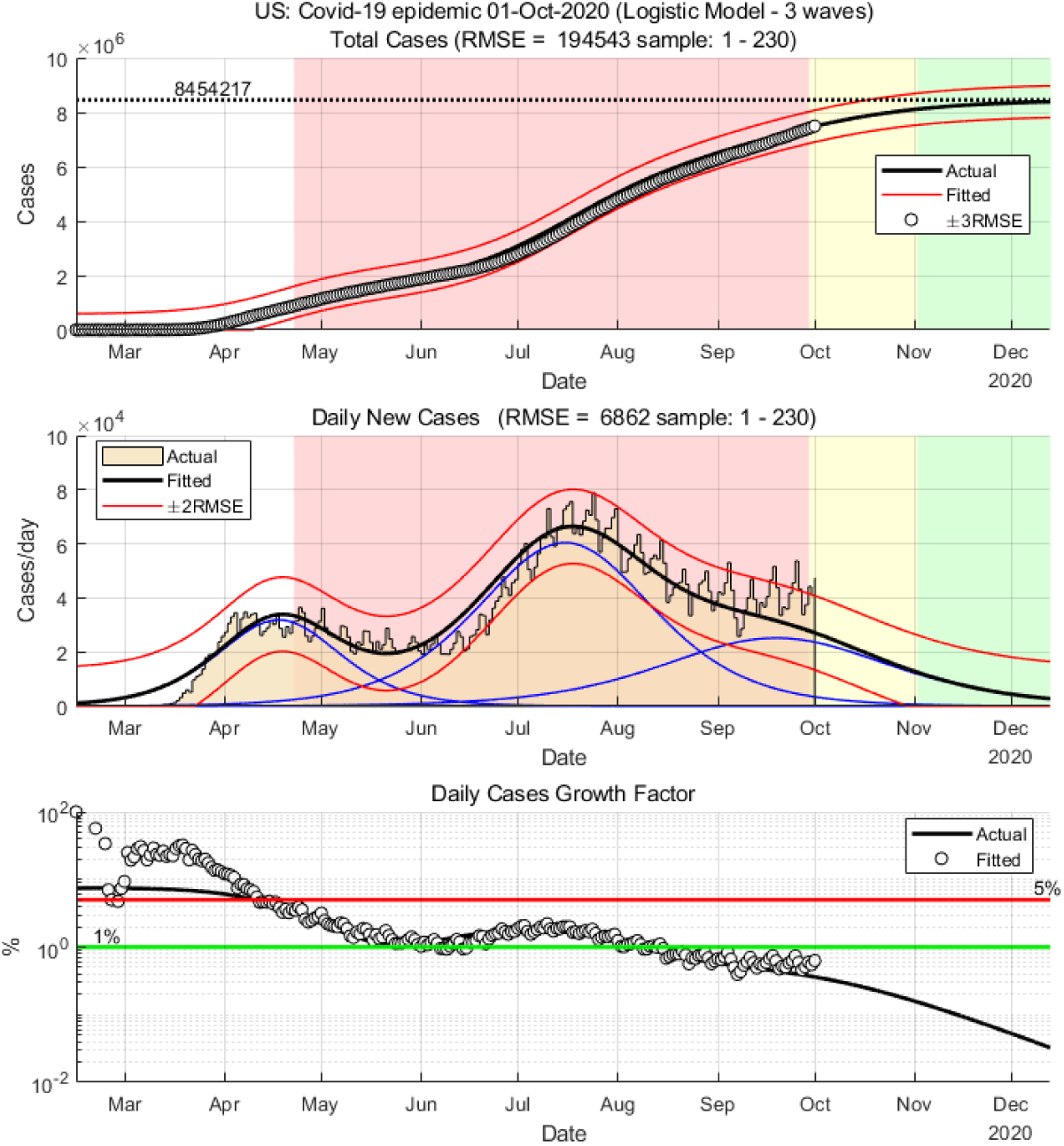
Current state and forecast of Covid 19 in the US.

**Figure 5a.**
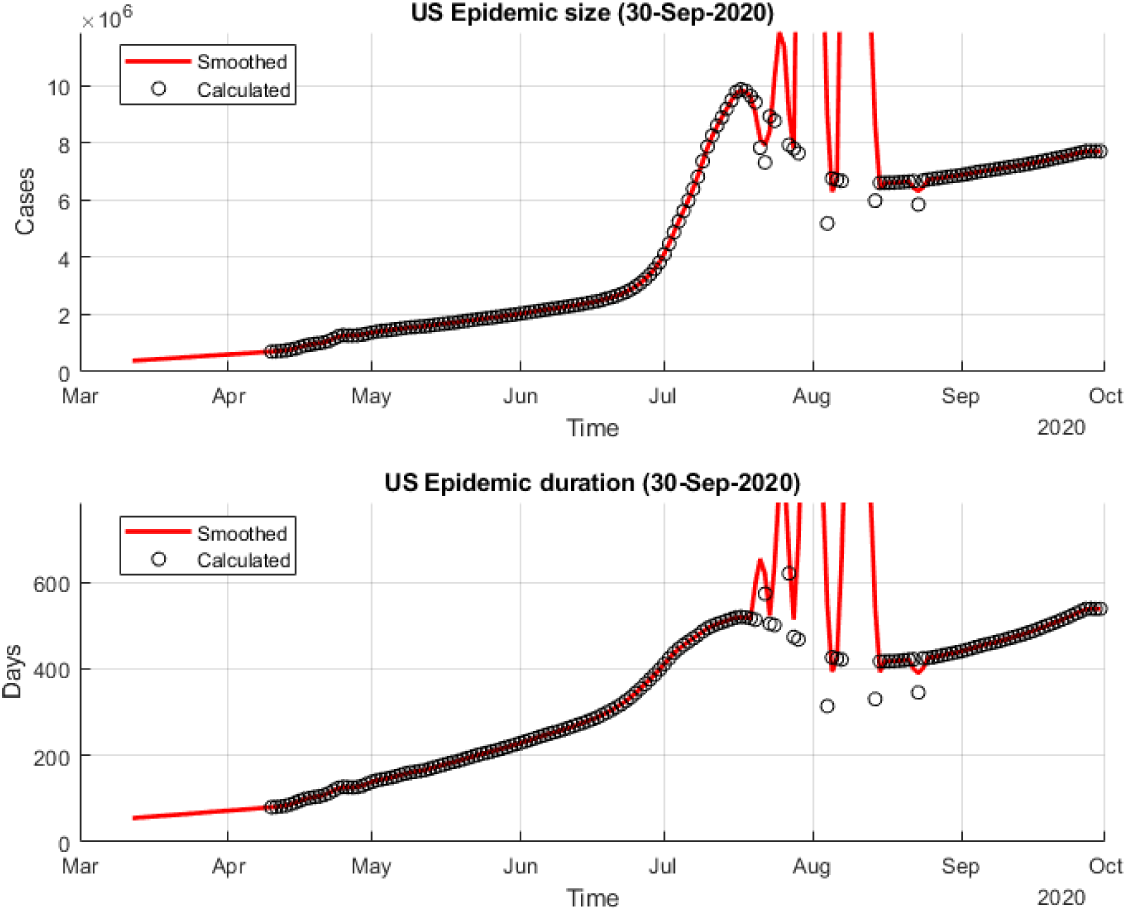
Daily-predicted epidemic size and duration for Covid 19 in the US.

**Figure 6a.**
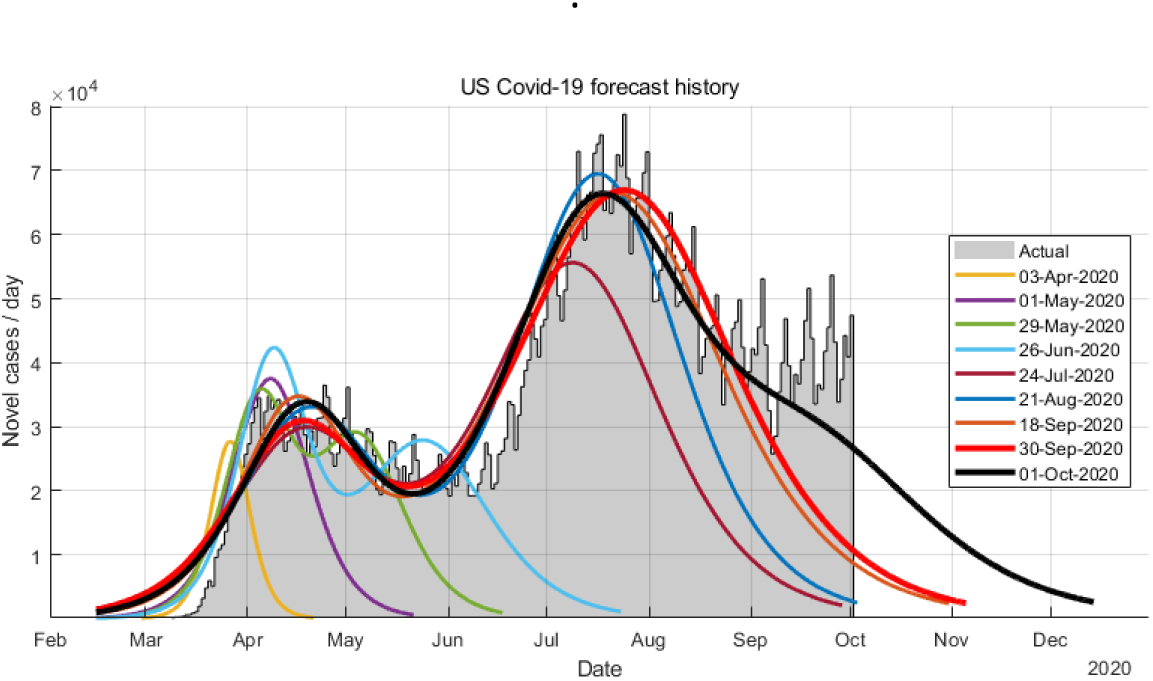
History of incidence forecasting for Covid 19 in USA.

## World

Current data suggest that the number of infected people worldwide could exceed 45 million. The epidemic can drag on into the spring of 2021. The current stagnation indicates the possibility of a third wave outbreak. The trend in the size and duration of the epidemic is still not stable, so these estimates are sure to change.

**Figure 7a.**
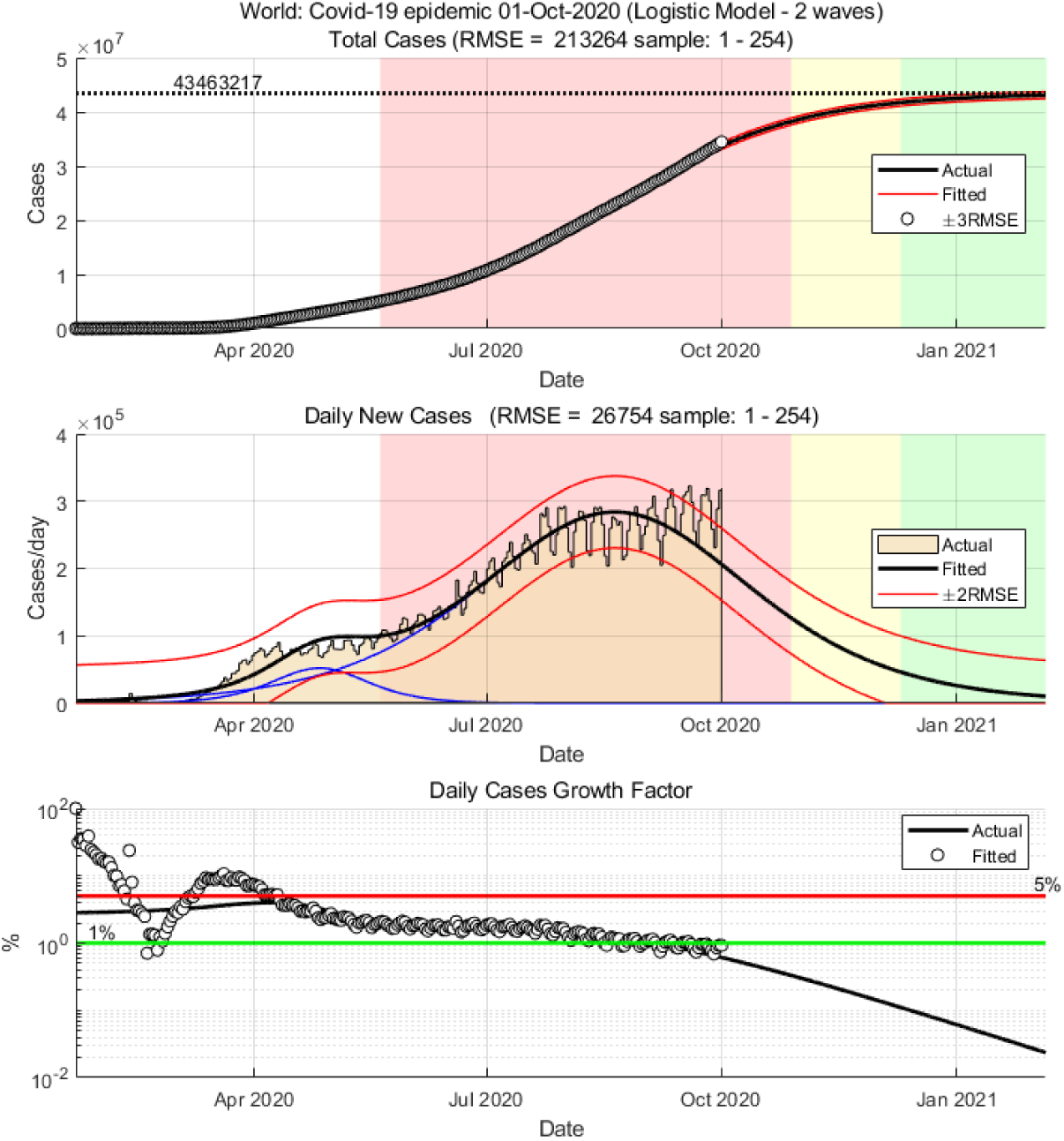
Situation and forecast of Corona 19 epidemic in the World.

**Figure 8a.**
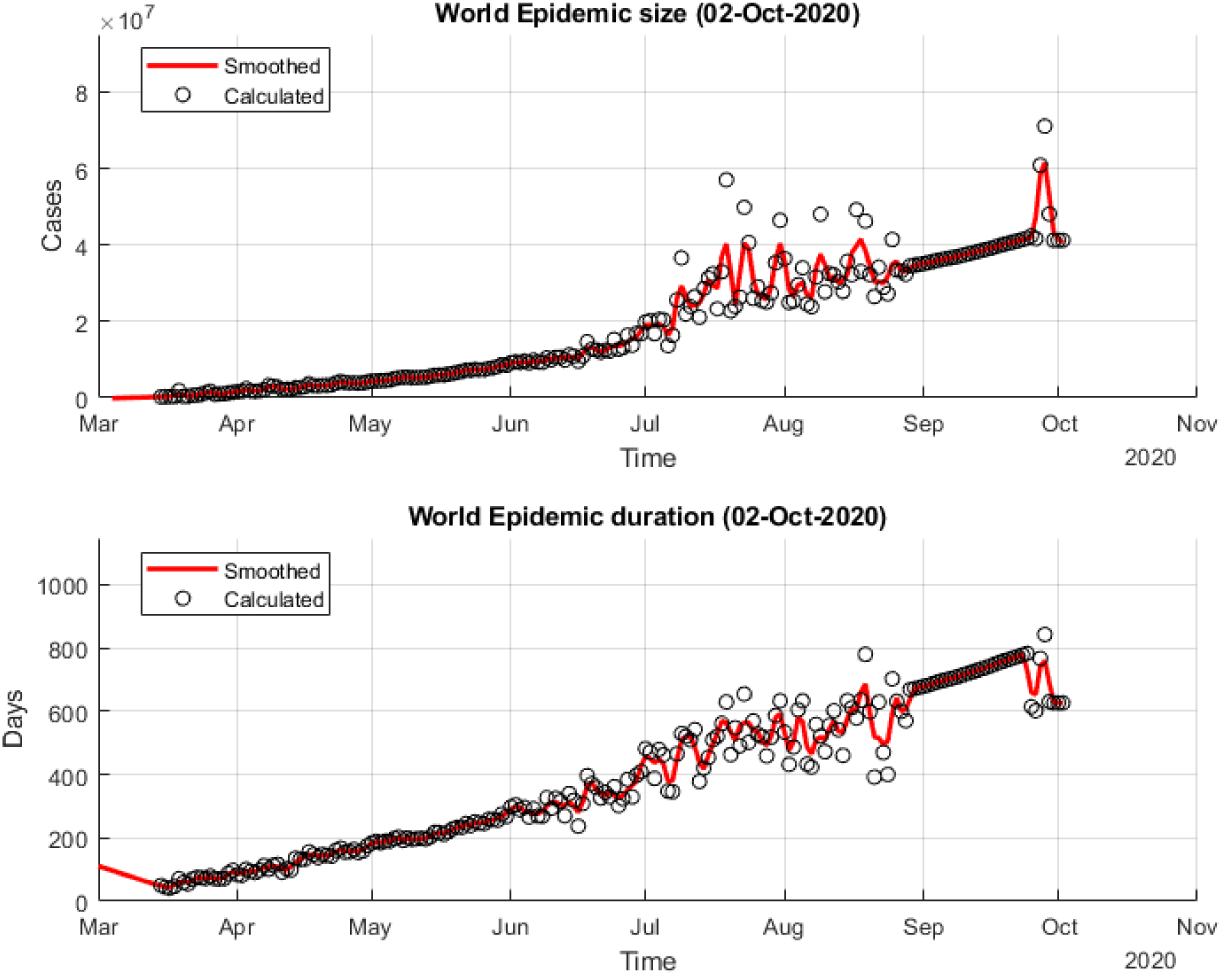
Daily-predicted epidemic size and duration for Covid 19 in the World.

**Figure 9a.**
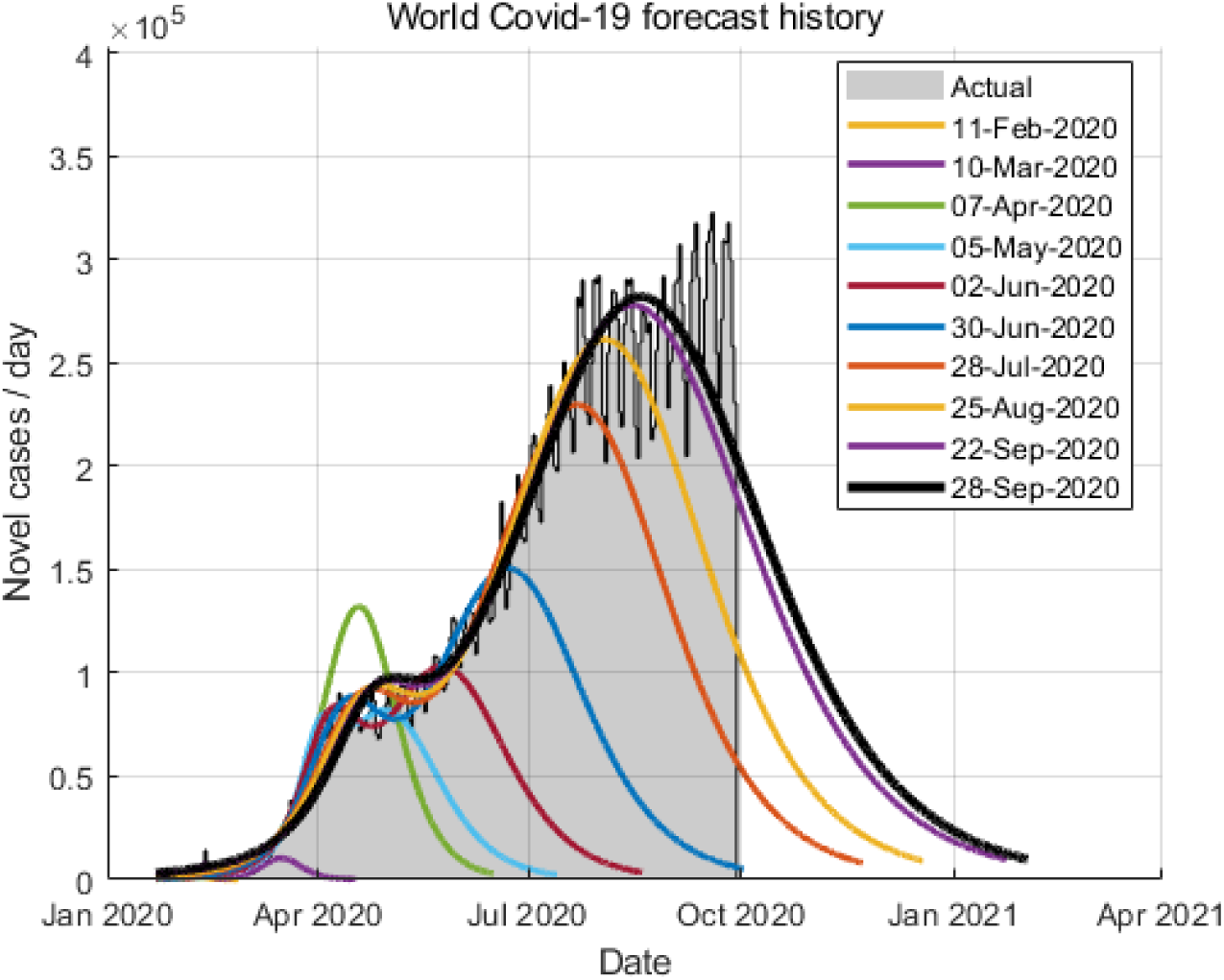
History of incidence forecasting for Covid 19 in World.

## Data Availability

The data used in this article are total confirmed cases, as are daily reported by Worldmeter.

https://www.worldometers.info/coronavirus/

## Footnotes

1 https://www.worldometers.info/coronavirus/

## References

Abdulrahman, I. K. (2020). SimCOVID: Open-Source Simulation Programs for the COVID-19 Outbreak. medRxiv, 2020.2004.2013.20063354. doi: 10.1101/2020.04.13.20063354

Agosto, A., & Giudici, P. (2020). A Poisson Autoregressive Model to Understand COVID-19 Contagion Dynamics. Risks, 8(3). doi: https://doi.org/10.3390/risks8030077

Anastassopoulou, C., Russo, L., Tsakris, A., & Siettos, C. (2020). Data-Based Analysis, Modelling and Forecasting of the COVID-19 outbreak. medRxiv, 2020.2002.2011.20022186. doi: 10.1101/2020.02.11.20022186

Bailey, N. T. J. (1975). The mathematical theory of infectious diseases and its applications (2nd ed.). London: Griffin.

Batista, M. (2020a). Estimation of the final size of the COVID-19 epidemic. medRxiv, 2020.2002.2016.20023606. doi: 10.1101/2020.02.16.20023606

Batista, M. (2020b). fitVirusXX, from https://www.mathworks.com/matlabcentral/fileexchange/76956-fitvirusxx

Bettencourt, L. M. A., & Ribeiro, R. M. (2008). Real Time Bayesian Estimation of the Epidemic Potential of Emerging Infectious Diseases. Plos One, 3(5).

Brauer, F. (2019a). Early estimates of epidemic final sizes. Journal of Biological Dynamics, 13(Sup1), 23–30. doi:10.1080/17513758.2018.1469792.

Brauer, F. (2019b). The Final Size of a Serious Epidemic. Bulletin of mathematical biology, 81(3), 869–877. doi: 10.1007/s11538-018-00549-x

Ceylan, Z. (2020). Estimation of COVID-19 prevalence in Italy, Spain, and France. Science of The Total Environment, 729, 138817. doi: https://doi.org/10.1016/j.scitotenv.2020.138817

Chowell, G., Ammon, C. E., Hengartner, N. W., & Hyman, J. M. (2006). Transmission dynamics of the great influenza pandemic of 1918 in Geneva, Switzerland: Assessing the effects of hypothetical interventions. Journal of Theoretical Biology, 241(2), 193–204.

Chowell, G., Tariq, A., & Hyman, J. M. (2019). A novel sub-epidemic modeling framework for short-term forecasting epidemic waves. Bmc Medicine, 17(1).

Daley, D. J., & Gani, J. M. (2001). Epidemic modelling an introduction ([Reprinted] ed.). Cambridge: Cambridge University Press.

Fisman D K. E., Tuite A.. (2014). Early Epidemic Dynamics of the West African 2014 Ebola Outbreak: Estimates Derived with a Simple Two-Parameter Model. PLOS Currents Outbreaks.. doi: 10.1371/currents.outbreaks.89c0d3783f36958d96ebbae97348d571

Frauenthal, J. C. (1980). Mathematical modeling in epidemiology. Berlin a.o.: Springer.

Giordano, G., Blanchini, F., Bruno, R., Colaneri, P., Di Filippo, A., Di Matteo, A., & Colaneri, M. (2020). Modelling the COVID-19 epidemic and implementation of population-wide interventions in Italy. Nature Medicine, 26(6), 855–860. doi: 10.1038/s41591-020-0883-7

He, S., Tang, S., & Rong, L. (2020). A discrete stochastic model of the COVID-19 outbreak: Forecast and control. Mathematical Biosciences and Engineering, 17(4), 2792–2804. doi: http://dx.doi.org/10.3934/mbe.2020153

He, S. B., Peng, Y. X., & Sun, K. H. (2020). SEIR modeling of the COVID-19 and its dynamics. Nonlinear Dynamics.

Hethcote, H. W. (2000). The mathematics of infectious diseases. Siam Review, 42(4), 599–653.

House, T., Ross, J. V., & Sirl, D. (2013). How big is an outbreak likely to be? Methods for epidemic final-size calculation. Proceedings of the Royal Society a-Mathematical Physical and Engineering Sciences, 469(2150).

Hsieh, Y. H., & Cheng, Y. S. (2006). Real-time forecast of multiphase outbreak. Emerging Infectious Diseases, 12(1), 122–127.

Keeling, M. J., & Rohani, P. (2008). Modeling infectious diseases in humans and animals. Princeton, N.J.: Princeton University Press.

Klepac, P., Kissler, S., & Gog, J. (2018). Contagion! The BBC Four Pandemic - The model behind the documentary. Epidemics, 24, 49–59.

Loli Piccolomini, E., & Zama, F. (2020). Monitoring Italian COVID-19 spread by a forced SEIRD model. PLoS ONE, 15(8). doi: https://doi.org/10.1371/journal.pone.0237417

Lopez, L. R., & Rodo, X. (2020). A modified SEIR model to predict the COVID-19 outbreak in Spain and Italy: simulating control scenarios and multi-scale epidemics. medRxiv, 2020.2003.2027.20045005. doi: 10.1101/2020.03.27.20045005

Maier, B. F., & Brockmann, D. (2020). Effective containment explains subexponential growth in recent confirmed COVID-19 cases in China. Science, 368(6492), 742-+.

Mbuvha R, & T, M. (2020). Bayesian inference of COVID-19 spreading rates in South Africa. PLoS ONE, 15(8). doi: https://doi.org/10.1371/journal.pone.0237126

Ming, W.-K., Huang, J., & Zhang, C. J. P. (2020). Breaking down of healthcare system: Mathematical modelling for controlling the novel coronavirus (2019-nCoV) outbreak in Wuhan, China. bioRxiv, 2020.2001.2027.922443. doi: 10.1101/2020.01.27.922443

Nesterov, Y. (2020). Online prediction of COVID19 dynamics. Belgian case study.. CORE Discussion Papers(22), 28.

Nesteruk, I. (2020). Statistics based predictions of coronavirus 2019-nCoV spreading in mainland China. medRxiv, 2020.2002.2012.20021931. doi: 10.1101/2020.02.12.20021931

Peng, L., Yang, W., Zhang, D., Zhuge, C., & Hong, L. (2020). Epidemic analysis of COVID-19 in China by dynamical modeling. medRxiv, 2020.2002.2016.20023465. doi: 10.1101/2020.02.16.20023465

Pongkitivanichkul, C., Samart, D., Tangphati, T., Koomhin, P., Pimton, P., Dam-O, P., … Channuie, P. (2020). Estimating the size of COVID-19 epidemic outbreak. Physica Scripta, 95(8), 085206. doi: 10.1088/1402-4896/ab9bdf

Roberts, D. H. (2020). A New Adaptive Logistic Model for Epidemics and the Resurgence of COVID-19 in the United States. medRxiv, 2020.2007.2017.20156109. doi: 10.1101/2020.07.17.20156109

Roda, W. C., Varughese, M. B., Han, D., & Li, M. Y. (2020). Why is it difficult to accurately predict the COVID-19 epidemic? Infectious Disease Modelling, 5, 271–281. doi: https://doi.org/10.1016/j.idm.2020.03.001

Singhal, A., Singh, P., Lall, B., & Joshi, S. D. (2020). Modeling and prediction of COVID-19 pandemic using Gaussian mixture model. Chaos, Solitons & Fractals, 138, 110023. doi: https://doi.org/10.1016/j.chaos.2020.110023

Tang, B., Wang, X., Li, Q., Bragazzi, N. L., Tang, S. Y., Xiao, Y. N., & Wu, J. H. (2020). Estimation of the Transmission Risk of the 2019-nCoV and Its Implication for Public Health Interventions. Journal of Clinical Medicine, 9(2).

Verity, R., Okell, L. C., & Dorigatti, I. (2020). Estimates of the severity of coronavirus disease 2019: a model-based analysis (vol 20, pg 669, 2020). Lancet Infectious Diseases, 20(6), E116–E116.

Wang, X. S., Wu, J. H., & Yang, Y. (2012). Richards model revisited: Validation by and application to infection dynamics. Journal of Theoretical Biology, 313, 12–19.

Wu, J. T., Leung, K., & Leung, G. M. (2020). Nowcasting and forecasting the potential domestic and international spread of the 2019-nCoV outbreak originating in Wuhan, China: a modelling study. The Lancet. doi: https://doi.org/10.1016/S0140-6736(20)30260-9

Yang, C. Y., & Wang, J. (2020). A mathematical model for the novel coronavirus epidemic in Wuhan, China. Mathematical Biosciences and Engineering, 17(3), 2708–2724.

Yang, W., Zhang, D., Peng, L., Zhuge, C., & Hong, L. (2020). Rational evaluation of various epidemic models based on the COVID-19 data of China. medRxiv, 2020.2003.2012.20034595. doi: 10.1101/2020.03.12.20034595

Zahiri, A., RafieeNasab, S., & Roohi, E. (2020). Prediction of Peak and Termination of Novel Coronavirus Covid-19 Epidemic in Iran. medRxiv, 2020.2003.2029.20046532. doi: 10.1101/2020.03.29.20046532

Zhan, C., Tse, C., Lai, Z., Hao, T., & Su, J. (2020). Prediction of COVID-19 spreading profiles in South Korea, Italy and Iran by data-driven coding. PLoS ONE 15(7). doi: https://doi.org/10.1371/journal.pone.0234763

Zou Y, Pan S, Zhao P, Han L, Wang X, Hemerik L, … W, v. d. W. (2020). Outbreak analysis with a logistic growth model shows COVID-19 suppression dynamics in China. PLoS ONE, 15(6). doi: https://doi.org/10.1371/journal.pone.0235247

